# Safety and immunogenicity of a single dose of a JN.1 variant COVID-19 vaccine in previously vaccinated adults: Primary analysis report of a phase 3 open-label trial

**DOI:** 10.1101/2025.08.20.25334074

**Authors:** Katia Alves, Alex Kouassi, Joy Nelson, Joyce S. Plested, Raj Kalkeri, Mingzhu Zhu, Shane Cloney-Clark, Zhaohui Cai, Ashika Ahmed, Irene McKnight, Raburn M. Mallory, Fernando Noriega

## Abstract

**Background:** We evaluated the immunogenicity and safety of a dose of NVX-CoV2705, a JN.1 subvariant SARS CoV-2 rS vaccine, in adults previously vaccinated with authorized/approved COVID-19 vaccines.

**Methods:** Study 2019nCoV-315 is a Phase 3, open-label, single-arm study to evaluate the safety and immunogenicity of a single dose of NVX-CoV2705 in previously COVID-19 vaccinated adult participants ≥18 years of age in the United States. Participants received one dose of NVX-CoV2705. The primary immunogenicity endpoint was the geometric mean titer (GMT) of serum neutralizing antibodies (nAbs) against the SARS-CoV-2 Omicron JN.1 variant at Day 28 following study vaccination and seroresponse rate (SRR) in ID_50_ titers for the JN.1 subvariant assessed at Day 28 following study vaccination. Primary safety endpoints include local and systemic solicited adverse events (AEs) up to day 6 post-vaccination, unsolicited AEs up to day 28, and treatment-related MAAEs, AESIs, and SAEs through Day 180. Exploratory endpoints also provided additional data for circulating variants.

**Results:** Between 14 October 2024 and 15 October 2024, 66 participants were screened and 60 enrolled. A total of 58 participants were included in the Per Protocol Analysis Set. GMTs increased from 138.6 (95% CI: 88.0–218.2) to 671.4 (95% CI: 4377.0–1031.6) from baseline to Day 28. In addition, GMTs increased ≥3.8□fold (GMFR) from baseline (Day 0) to Day 28 for currently circulating or emerging Omicron subvariants JN.1, LP.8.1, KP.2, KP.3, KP.3.1.1, MC.1, XEC, MC.10.1, LF.7, LF.7.2.1, LF.7.7.2, NB.1.8.1, XFC, and XFG, with an acceptable safety profile after single dose vaccination.

**Discussion:** A single dose of NVX-CoV2705 induced a rapid and robust anti–SARS-CoV-2 immune response against the Omicron JN.1 and other circulating variants and had an acceptable safety profile.

**ClinicalTrials.gov identifier:** NCT06409663

## Introduction

Since 2020, severe acute respiratory syndrome coronavirus 2 (SARS-CoV-2), the virus that causes coronavirus disease 2019 (COVID-19), has been spreading and changing globally. It has been over 5 years since the advent of COVID-19, and the disease has affected over 775 million people.^1^ Although the number of COVID-19 related deaths is substantially lower than what was seen during the pandemic, the SARS-CoV-2 virus still remains a substantial burden, particularly in older adults and immunocompromised patients.^2,3^ The virus has continued to spread and evolve globally due to selective pressures such as host immune responses and mutations that increase transmissibility. These emerging SARS-CoV-2 variants, including the Omicron lineages and its descendant strains (JN.1, KP.2, KP.3, and XEC), contain mutations that increase resistance to both vaccine-induced and natural immunity^4,5^ and are still infecting people around the world.^6^ At the June 5^th^, 2024, meeting of FDA’s Vaccines and Related Biological Products Advisory Committee (VRBPAC), members voted that vaccines for the 2024-2025 vaccination campaign be based on JN.1.^7^

NVX-CoV2373 (Novavax) is a SARS-CoV-2 vaccine that contains 5 μg recombinant nanoparticle spike protein (SARS-CoV-2 rS) and 50 μg Matrix-M adjuvant^TM^.^8^ Novavax has developed this vaccine for the proposed indication of active immunization for the prevention of COVID-19 caused by SARS□CoV-2 in individuals 12 years of age and older. The original, ancestral Wuhan strain-based vaccine (Nuvaxovid^®^) was updated for the 2024 -2025 season to a monovalent Omicron JN.1 subvariant vaccine (NVX-CoV2705). This formulation aligns with recommendations issued by VRBPAC, the World Health Organization (WHO), European Medicines Agency (EMA), and the International Coalition of Medicines Regulatory Authorities (ICMRA) for vaccine composition for the 2024-2025 vaccination season.^9,10^

Here we present results describing the immunogenicity at Day 28 and protocol-defined Day 180 safety results for NVX-CoV2705, a monovalent Omicron JN.1 subvariant vaccine.

## Methods

### Study design and participants

This was a phase 3, single-arm, open-label trial conducted at two sites in the United States (ClinicalTrials.gov identifier NCT06409663). In this study, participants received a single intramuscular dose of NVX-CoV2705. Participants were medically stable male and nonpregnant females who were ≥18 years of age and previously vaccinated with authorized/approved COVID-19 vaccines (Figure 1). Participants’ last COVID-19 vaccination must have been administered ≥6 months prior to study vaccination. Key exclusion criteria were pregnancy; receiving any other vaccine within 30 days prior to study vaccination or planning to receive any other vaccine within 28 days after study vaccination (influenza vaccines could be administered up to 14 days before study vaccination); any known history of allergies or anaphylaxis to products contained in the investigational product in the participant’s lifetime; and known history of myocarditis or pericarditis in the participant’s lifetime.

**Figure 1.**
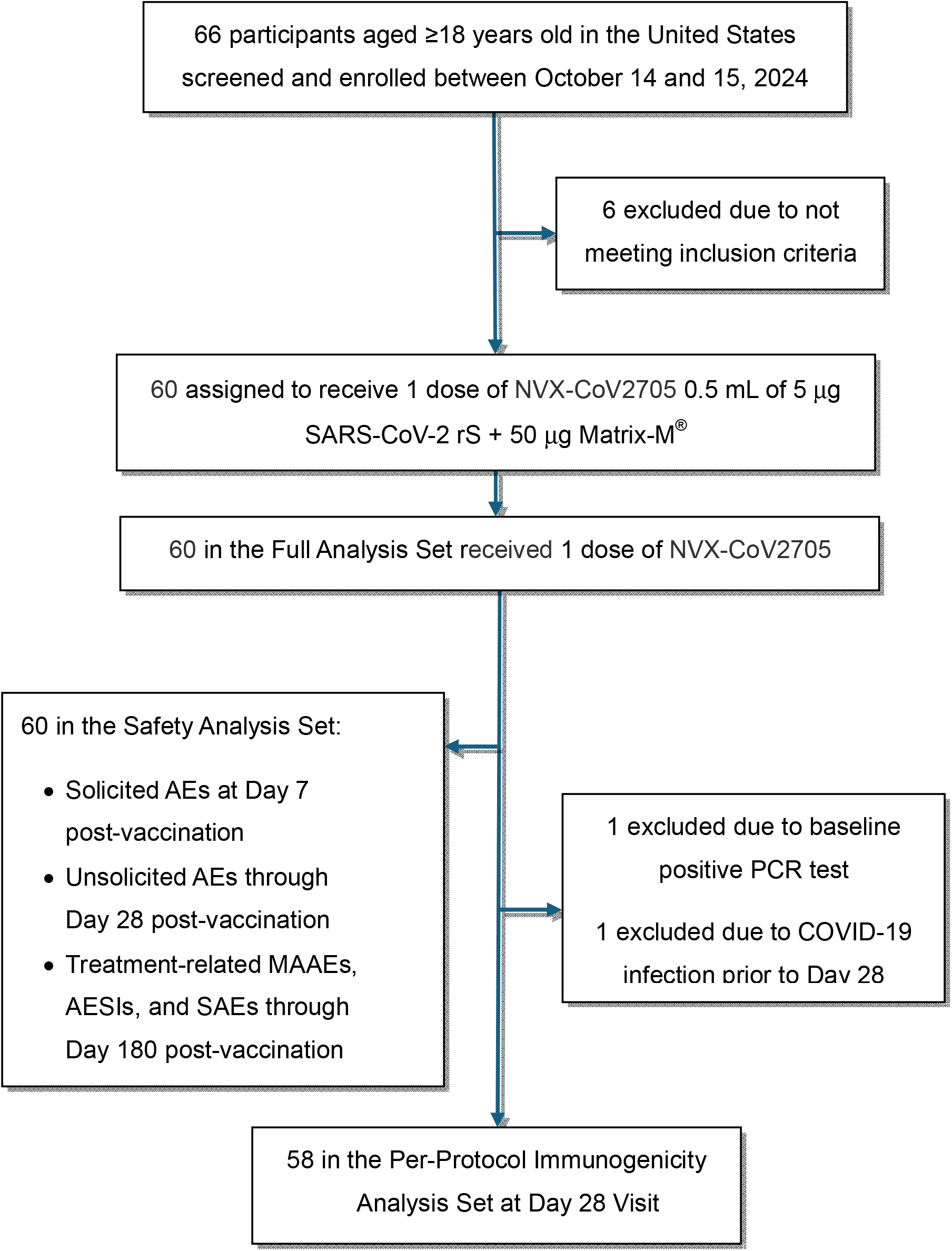
Consort diagram.

The study commenced on October 14, 2024, and was completed on May 9, 2025. The protocol and amendments were approved by an institutional review board. The study was conducted in accordance with the International Conference on Harmonisation Good Clinical Practice Guidelines, the Declaration of Helsinki ethical principles, and all other applicable laws and regulations. Written informed consent was obtained from all study participants before enrollment.

### Objectives and endpoints

The primary immunogenicity objective was to describe the immune response to NVX-CoV2705. The geometric mean titer (GMT) of serum neutralizing antibodies (nAbs) against the SARS-CoV-2 Omicron JN.1 variant at Day 28 following study vaccination and seroresponse rate (SRR) in ID_50_ titers for the JN.1 subvariant assessed at Day 28 following study vaccination were described.

The primary safety objective was to describe the safety and tolerability profile of NVX□CoV2705. The primary safety endpoints were incidence, duration, and severity of solicited local and systemic AEs for 7 days following vaccination; incidence, severity, and relationship of any unsolicited AEs through 28 days after vaccination; and incidence and severity of treatment-related MAAEs, AESIs (predefined list, including PIMMCs, myocarditis and/or pericarditis), and SAEs through Day 180. AEs were graded by the investigator using the FDA Toxicity Grading Scale.^11^ Participants utilized a paper diary card to record reactogenicity following vaccination on Day 0 and then daily for an additional 6 days after vaccination. All diary entries were reviewed and assessed by the investigator and paper diary cards were collected at the Day 6 in-person Visit. Participants were tested for SARS-CoV-2 infection on Day 0.

The secondary immunogenicity endpoint was to describe serum anti-S IgG antibody responses against the Omicron JN.1 variant at Day 28 following study vaccination compared to baseline.

An exploratory endpoint analyzed the cross-reactivity of neutralizing antibody responses of NVX-CoV2705 against multiple SARS-CoV-2 pseudovirus strains using both validated (ancestral Wuhan strain and Omicron BA.5, XBB.1.5, JN.1, KP.2, and KP.3 subvariants) and fit-for-purpose (Omicron KP.3.1.1, XEC, MC.1, LP.8.1, LF.7.2.1, LF.7.7.2, NB.1.8.1, XFC, and XFG subvariants) assays.

### Statistical analysis

The immunogenicity population and safety population included participants who were eligible for the study and who received the study vaccine. The per protocol set (PPS) included all participants who received the vaccine, were PCR negative at baseline, and had no major protocol violations. The two primary immunogenicity endpoints and the secondary and exploratory immunogenicity endpoints were analyzed using the PPS and were analyzed using descriptive statistics. Additional descriptive analyses of GMT and SRR were performed separately by age group (18–54 years of age, ≥55 years of age) and baseline anti-nucleocapsid (anti-N) status. Participants with missing values for these subgroup variables were not included in the subgroup analyses. The durations of solicited local and systemic AEs and unsolicited AEs after vaccination within the 7 Day diary period and entire safety follow-up period were also summarized. All statistical analyses were completed using SAS^®^ version 9.4 or higher.

## Results

### Participants

The study was conducted at two sites in the United States. From October 14, 2024, to October 15, 2024, 66 participants were screened (Figure 1), and 60 participants were enrolled in the study and received a single dose of NVX-CoV2705 (0.5 mL containing 5 μg SARS-CoV-2 rS plus 50 μg Matrix-M adjuvant) via a deltoid intramuscular injection (Figure 1, CONSORT diagram). Immunogenicity was assessed at Days 0 and 28. The data cutoff for all analyses was May 9, 2025. Safety data from Day 0 to Day 28 were assessed; safety data were also assessed at Day 90 and Day 180. There were six screen failures, all of which were attributed to failure to meet the inclusion/exclusion criteria. The Full Analysis Set (FAS) and Safety Analysis Set comprised all enrolled participants. The PP Analysis Set at the Day 28 visit comprised 58 (96.7%) participants, with one participant excluded due to a positive PCR result at baseline and one excluded due to a COVID-19 infection prior to the Day 28 visit.

Participant demographics and characteristics for the Safety Analysis Set are shown in Table 1. The median age (range) of participants in the PP Analysis Set was 57.0 years (21–82 years), with the majority of participants ≥55 years of age (58.3%). The majority of participants were female (63.3%), White (75.0%), and not of Hispanic or Latino origin (76.7%); Black or African American participants accounted for 20.0% of participants. Most participants were either overweight or obese (80.0%) and anti–N-positive (88.3%), and all participants included in the PP Analysis Set were PCR negative at baseline. One additional participant was excluded from the PP Analysis Set due to a COVID-19 infection prior to the Day 28 visit. All participants had previously received an authorized or approved COVID-19 vaccine, though the variant of the prior COVID-19 vaccine was largely unknown. The median time (range) between the last prior COVID-19 vaccine and study vaccination was 676 days (251 to 1456 days).

**Table 1.** Demographics and baseline characteristics of 2019nCoV-315 study participants.

|  | **JN.1 Subvariant SARS-CoV-2 rS Vaccine**<br>**(Safety Analysis Set), N= 60** |
| --- | --- |
| **Age** | Median (SD): 57.0 (13.62) |
| **Female, n (%)** | 38 (63.3) |
| **Race/Ethnicity, n (%)** |  |
| White | 45 (75.0) |
| Black | 12 (20.0) |
| Asian | 1 (1.7) |
| Mixed origin | 1 (1.7) |
| American Indian or Alaskan Native | 1 (1.7) |
| Not Hispanic or Latino | 46 (76.7) |
| **BMI (kg/m<sup>2</sup>), n (%)** |  |
| Underweight (<18) | 1 (1.7) |
| Normal (18 – 24.9) | 11 (18.3) |
| Overweight (25 – 29.9) | 20 (33.3) |
| Obese (>30) | 28 (46.7) |
| **Prior COVID-19 vaccinations, n (%)** |  |
| Moderna | 22 (36.7) |
| Pfizer | 21 (35.0) |
| Mixed | 16 (26.7) |
| Other | 1 (1.7) |

### Immunogenicity

The first primary endpoint was to describe the nAb response against the JN.1 pseudovirus following a single dose of NVX-CoV2705. GMTs increased from 138.6 (95% CI: 88.0, 218.2) to 671.4 (95% CI: 4377.0–1031.6) from baseline to Day 28 (Table 2). The GMFR was 4.5, with a two-sided 95% CI of 3.1, 6.5. The second primary endpoint was to describe the SRR for a single dose of NVX-CoV2705. The SRR following a single dose of NVX-CoV2705 was 37.5%, with a two-sided 95% CI (24.9–51.5%).

**Table 2.** Immunogenicity results of pseudovirus neutralizing titers (per-protocol set)

|  | **All Ages**<br>**N=59** |  | **Younger Adults**<br>**≥18–54 years of age**<br>**N=25** |  | **Older Adults**<br>**≥55 years of age**<br>**N=33** |  |
| --- | --- | --- | --- | --- | --- | --- |
| Baseline GMT (ID<sub>50</sub>),<br>95% CI (n1) | n=58 | 138.6<br>(88.0–218.2) | n=25 | 130.7<br>(66.2–257.9) | n=33 | 144.8<br>(776.2–275.1) |
| Day 28 GMT (ID<sub>50</sub>),<br>95% CI (n1) | n=56 | 671.4<br>(437.0–1031.6) | n=24 | 430.1<br>(213.2–867.5) | n=32 | 937.8<br>(546.3–<br>1609.7) |
| GMFR from baseline<br>to Day 28, 95% CI<br>(n2) | n=56 | 4.5<br>(3.1–6.5) | n=24 | 3.0<br>(1.8–5.1) | n=32 | 6.0<br>(3.7–10.0) |
| SRR from baseline to<br>Day 28 (%) (n3) | n=21 | 37.5<br>(24.9–51.5) | n=4 | 16.7<br>(4.7–37.4) | n=17 | 53.1<br>(344.7–70.9) |

GMT, geometric mean titers; GMFR, geometric mean fold rise; N, total number of participants in the analysis set; n, number of participants within each visit with non-missing data for this strain; SRR=, seroconversion rate

For the secondary endpoint of serum anti-S IgG levels compared to baseline, anti-S IgG antibody GMEUs against the JN.1 protein increased from 12491.9 (95% CI: 9548.1–16,343.4) EU/mL at baseline to 31129.0 (95% CI: 24,453453.6–39,626.5) EU/mL at Day 28. The adjusted GMFR was 2.4, with a two-sided 95% CI (1.9–2.9). The SRR on Day 28 was 19.6% (95% CI: 10.2–32 2.4%).

When neutralizing antibody responses to NVX-CoV2705 were measured against various strains of COVID-19, GMTs increased GMTs (ID_50_) increased ≥3.8□fold (GMFR) from baseline (Day 0) to Day 28 for currently circulating or emerging Omicron subvariants JN.1, LP.8.1, KP.2, KP.3, KP.3.1.1, MC.1, XEC, MC.10.1, LF.7, LF.7.2.1, LF.7.7.2, NB.1.8.1, XFC, and XFG (Figure 2).

**Figure 2.**
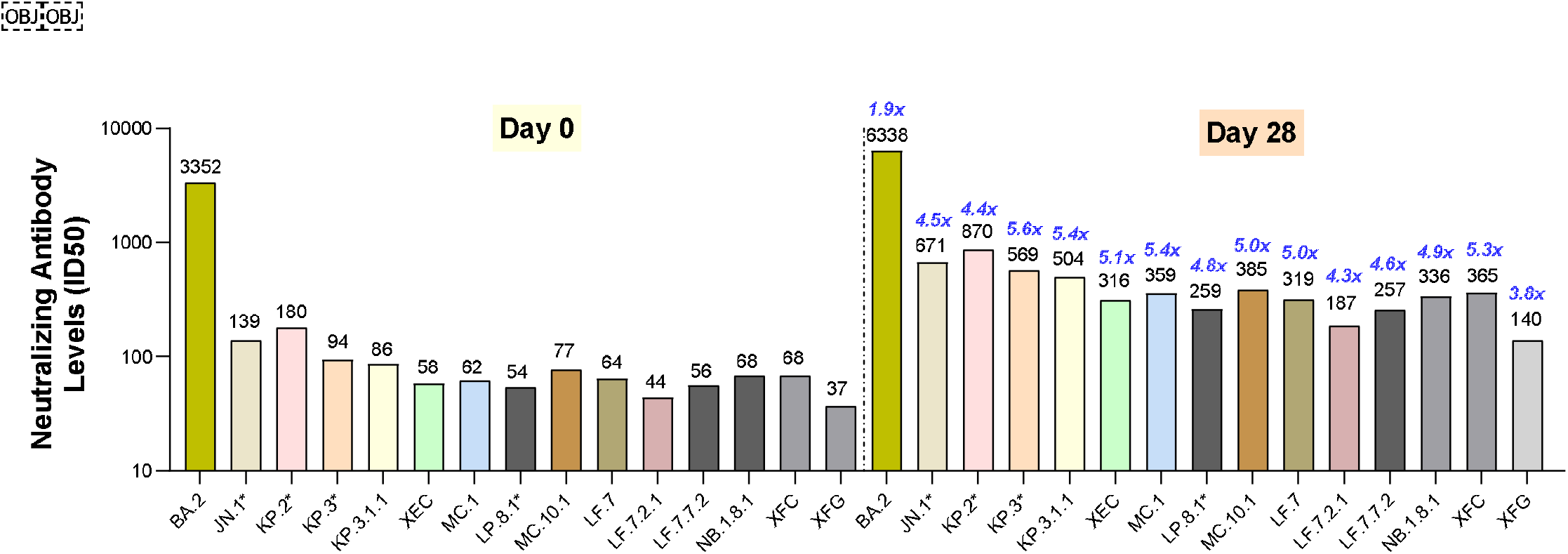
Immune responses to past and currently circulating strains post-JN.1 booster (Pseudovirus Neutralizing Titers). Note: ^*^ signifies the validated strains.

### Safety outcomes and assessments

Solicited local TEAEs were reported in 35 (58.3%) participants within 7 days following single dose vaccination with NVX-CoV2705. The overall incidence of solicited local TEAEs was higher in participants 18–54 years of age (64.0%) than in participants ≥55 years of age (54.3%) (Figure 3A). Most of them were grade 1 or grade 2 in severity, with 2 (3.3%) participants reporting grade 3 events of pain and tenderness and no participant reporting a grade 4 event (Table 3). Of note, all events of grade 3 and higher were reported in participants 18–54 years of age. Tenderness and pain of any grade were the most frequent (incidence >20%) solicited local injection site TEAEs, 45.7% and 42.9% in participants aged ≥55 years, respectively, and 64.0% and 48.0% in participants aged 18–54 years, respectively. The incidence of pain and tenderness of grade 3 and higher in participants aged 15–54 years was 8.0% for both symptoms, and none were reported by participants aged ≥55 years (Figure 3A). Median durations were 2 days for pain and tenderness, and 1 day for redness and swelling.

**Table 3.** Summary of unsolicited TEAEs.

| **Description** | **Participants with AEs**<br>**(Safety Set), N=60, n (%)** |
| --- | --- |
| Any TEAE | 10 (16.7) |
| Related TEAE | 1 (1.7) |
| Any MAAE | 8 (13.3) |
| Treatment-related MAAE | 0 |
| Serious AE | 1 (1.7) |
| Any Severe AEs | 2 (3.3) |
| Any AESI | 0 |
| AE leading to study discontinuation | 0 |
| Deaths | 0 |

AE, adverse event; MAAE, medically attended adverse event; TEAE, treatment-emergent adverse event; AESI, adverse event of special interest

**Figure 3A.**
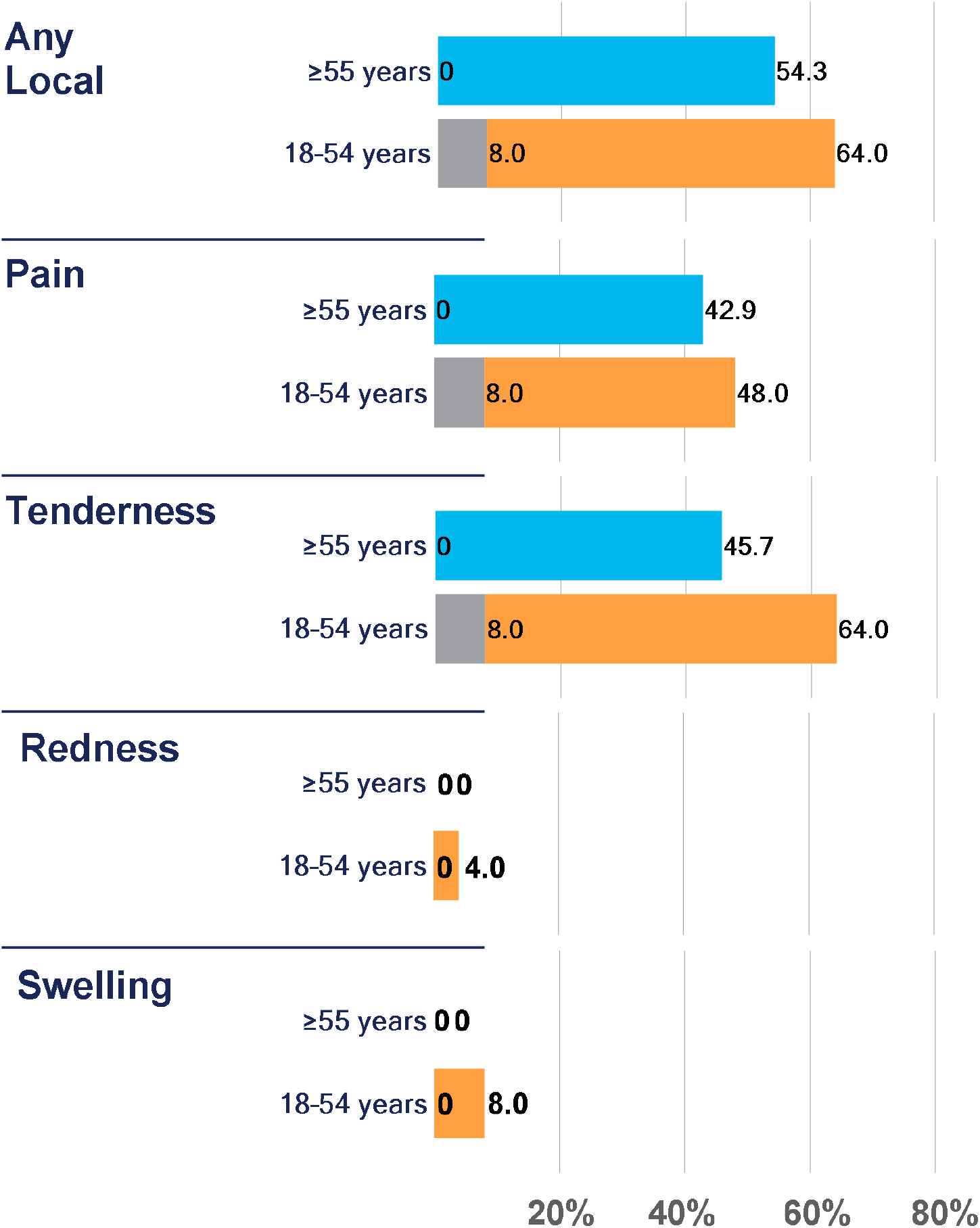
Solicited Local Injection Site TEAEs: Local Reactogenicity by Grade.

Solicited systemic TEAEs were reported in 35 (58.3%) participants within 7 days following single dose vaccination with NVX-CoV2705. The overall incidence of solicited systemic TEAEs was higher in participants aged 18–54 years (76.0%) than in participants aged ≥55 years (45.7%) (Figure 3B). Most of them were grade 1 or grade 2 in severity. Of note, any systemic TEAEs of grade 3 and higher occurred in 12% participants aged 15–54 years compared to 2.9% in participants aged ≥55 years. Any-grade fatigue/malaise (18–54 years old: 44.0%; ≥55 years old: 20.0%), malaise (18–54 years old: 24.0%; ≥55 years old: 11.4%), headache (32.0% and 28.6%, respectively), muscle pain (52.0% and 28.6%, respectively), and fatigue (36.0% and 20.0%, respectively) were most commonly reported. No grade 4 events occurred. Of note, solicited systemic TEAEs of grade 3 and higher were reported only in participants aged 18–54 years; fatigue/malaise (18–54 years old: 12.0%; ≥55 years old: 0.0%), headache (4.0% and 0.0%, respectively), fatigue (8.0% and 0.0%, respectively), malaise (8.0% and 0.0%, respectively), and joint pain (4.0% and 0.0%., respectively). Both age groups reported muscle pain of grade 3 and higher (18–54 years old: 8.0%; ≥55 years old: 2.9%), and none reported nausea/vomiting of grade 3 and higher (Figure 3B). Median durations were 2.5 days for malaise; 2 days for joint pain; 1.5 days for fatigue and nausea/vomiting; and 1 day for muscle pain and headache.

**Figure 3B.**
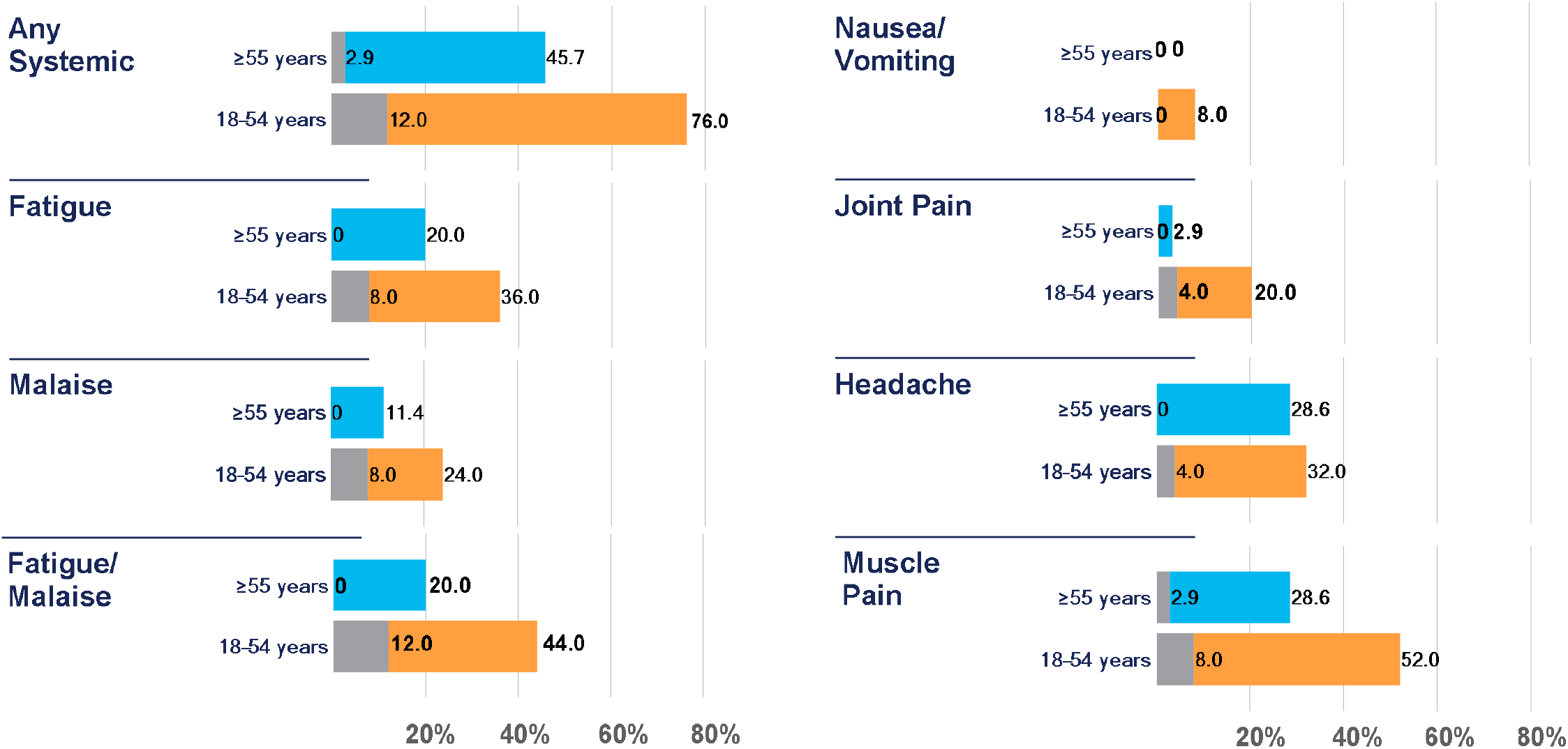
Solicited Systemic Injection Site TEAEs: Systemic Reactogenicity.

Unsolicited TEAEs were reported by 10 (16.7%) participants through 28 days after single dose vaccination with NVX-CoV2705 (Table 3). The most frequent unsolicited TEAEs (incidence >5%) were from the SOC Infections and Infestations, occurring in 4 (6.7%) participants. Contusion and skin abrasion each reported in 2 (3.3%) participants were the most frequent unsolicited TEAEs. There were 2 (3.3%) severe AEs, and 1(1.7%) SAE assessed as not related to the vaccine. No AESIs, myocarditis/ pericarditis, treatment-related MAAEs, AEs leading to study discontinuation, or deaths were reported.

## Discussion

The overwhelming majority of the US population, has received a vaccination for COVID-19, been infected with COVID-19, or both.^12,13^ The latest estimates indicate that 81.4% of the US population has received at least one dose of a COVID-19 vaccination,^12^ and a study of US blood donors reported that the combined seroprevalence from infection or vaccination reached 94.7% by December 2021.^13^ The objective of this study was to collect clinical data on the selected strain to be submitted as a post-authorization commitment to the US FDA and to gather data about the extent to which the selected strain was able to induce cross-neutralizing antibodies to strains that began to circulate subsequent to the aforementioned 2024 VRBPAC recommendation.

The present study aimed to investigate the safety and immunogenicity of an updated Novavax SARS-CoV-2 rS adjuvanted with Matrix-M based on the recommendation made by the US VRBPAC. This study was conducted in previously COVID-19 vaccinated participants ≥ 18 years of age and employed a JN.1 subvariant vaccine.

A total of 60 participants received a single dose of NVX-CoV2705 on Day 0 and remained on study for immunogenicity until Day 28 and safety data collection up to 180 days post-vaccination.

Both primary endpoints were described, as measured by pseudovirus neutralization (ID_50_) GMTs for the JN.1 subvariant at Day 28 following study vaccination and SRRs in ID_50_ titers for the JN.1 subvariant assessed at Day 28 following study vaccination. The descriptive results for neutralizing GMTs for the JN.1 subvariant levels are consistent with the pattern observed in previous vaccines targeting different COVID-19 variants. A robust increase in neutralizing antibody titers was noted, with some variability seen by age. Older adults (≥ 55 years of age) displayed a greater increase in pseudovirus neutralizing titers than participants 18 to 54 years of age,. This is contrary to what has been reported in previous trials, in which younger participants displayed a more robust response.^14-16^ This may be attributed to the small sample sizes for this subgroup analysis, which resulted in broad and overlapping confidence factors for the GMT estimates; however, other factors such as enhanced immune imprinting from prior strains in younger participants could also play a role. A planned study evaluating the immunogenicity and safety of the JN.1 variant vaccine in adolescents and adults 12 through 64 years of age who have risk factors for severe COVID-19 and in adults 65 years of age and older may indicate whether this was a chance finding.

The secondary endpoint was assessed and, on Day 28, anti-S IgG antibody GMEUs against the JN.1 protein increased almost 3.0-fold from baseline.

For the exploratory endpoint, pseudovirus neutralizing GMTs increased ≥4.3-fold for current circulating or emerging Omicron subvariants at Day 28, while the increase was less robust at 1.5 and <3-fold for the ancestral Wuhan strain and prior circulating Omicron subvariants BA.5 and XBB 1.5.

No new safety signals were identified in the analysis of safety and reactogenicity. The demographics for this study showed good proportional representation, although our sample size was small. Additionally, this was an open label study that lacked a comparison group. Importantly, safety and reactogenicity of NVX-CoV2705 were consistent with that of the prototype vaccine, NVX-CoV2373.^14-16^

## Conclusions

Single dose administration of NVX-CoV2705 in previously authorized/approved COVID-19 vaccinated participants showed robust immunogenicity against the JN.1 variant and related variants derived from the JN.1 strain. NVX-CoV2705 was well tolerated, with an acceptable profile, following single dose administration. At the time of this report, the Novavax COVID-19 vaccine is authorized for use in adults 65 years and older and for individuals 12 through 64 years who have at least one underlying condition that puts them at high risk for severe outcomes from COVID□19. The results presented herein demonstrate that an antigen-updated single dose of this vaccine is safe and immunogenic.

## Data Availability

Study information is available online at https://www.clinicaltrials.gov/study/NCT06409663. Requests submitted to the corresponding author will be considered upon publication of this article and de-identified participant data, related to results reported here, may be provided.

## Author contributions

KA, RMM, and FN were involved in the study design and conducted the data analysis and interpretation. KA, JSP, RK, MZ, SCC, ZC, AA, and FN were involved in data collection. JN and SCC provided project management. AK performed the statistical analyses. KA, JSP, RK, MZ, SCC, ZC, AA, and FN have directly accessed and verified the data in this manuscript. IM wrote the original draft of this manuscript. KA, JSP, RK, MZ, SCC, ZC, AA, IM, RMM, and FN were involved in data interpretation and reviewed, commented on, and approved this manuscript prior to submission for publication. The authors accept accountabilities for all aspects of the work, ensuring questions related to accuracy or integrity are investigated and resolved. All authors had full access to all the data in the study and had final responsibility for the decision to submit for publication.

## Ethical considerations

The study protocols for all sites in the 2019nCoV-315 study was approved by Advarra, and the study was conducted according to the principles of the International Conference on Harmonisation Good Clinical Practice Guideline, adopting the principles of the Declaration of Helsinki, as well as all applicable national, state, and local laws and regulations. All participants provided written informed consent.

## References

1. World Health Organization. Number of COVID-19 cases reported to WHO (cumulative total). Available at http://data.who.int/dashboards/COVID-19/cases. Accessed 11 March 2025.

2. Kaku Y, Uriu K, Kosugi Y, et al. Virological characteristics of the SARS-CoV-2 KP.2. Lancet Infect Dis. 2024;24:e416.

3. Kaku Y, Okumuru K, Kawakubo S, et al. Virological characteristics of the SARS-CoV-2 XEC variant. Lancet Infect Dis. 2024;24:e736.

4. Markov PV, Ghafari M, Beer M, et al. The evolution of SARS-CoV-2. Nat Rev Microbiol. 2023;21(6):361–379.

5. Lin DY, D. Y, Xu Y, et al. Durability of XBBB.1.5 vaccines against omicron subvariants. N Engl J Med. 2024;390(22): 2124–2127.

6. Seneviranthe TH, Wekking D, Swain JWR, Solinas C, DeSilva P. COVID-19: From emerging variants to vaccination. Cytokine Growth Factor Rev. 2024;76:127–141.

7. US Food and Drug Administration. Vaccines and Related Biological Products Advisory Committee June 5, 2024 Meeting Announcement. Available at: https://www.fda.gov/advisory-committees/advisory-committee-calendar/vaccines-and-related-biological-products-advisory-committee-june-5-2024-meeting-announcement. Accessed 19 March 2025.

8. Keech C, Albert G, Cho A, et al. Phase1–2 trial of a SARS-CoV-2 recombinant spike protein nanoparticle vaccine. N Engl J Med 2020; 383:2320–32. 10.1056/NEJMoa2026920.

9. United States Food and Drug Administration. Recommendation for the 2023-2024 formula of COVID-19 vaccine in the US. Accessed on July 26, 2023. Available at: https://www.fda.gov/media/169591/download.

10. World Health Organization. Statement on the antigen composition of COVID-19 vaccines. Accessed on July 26, 2023. Available at: https://www.who.int/news/item/18-05-2023-statement-on-the-antigen-composition-of-covid-19-vaccines.

11. U.S. Food and Drug Administration. Guidance for industry: toxicity grading for healthy adult and adolescent volunteers enrolled in preventative vaccine clinical trials. 2007[Cited:23-02-2023]; Available from: https://www.fda.gov/media/73679/download.

12. Centers for Disease Control and Prevention. National Center for Immunization and Respiratory Diseases (NCIRD). The changing threat of COVID-19. February 23, 2024. Available at:https://www.cdc.gov/ncird/whats-new/changing-threat-covid-19.html. Accessed 24 April 2025.

13. Jones JM, Opsomer JD, Stone M, Benoit T, Ferg RA, Stramer SL, Busch MP. Updated US infection-and vaccine-induced SARS-CoV-2 seroprevalence estimates based on blood donations, July 2020-December 2021. JAMA. 2022;328(3):298–301.

14. Dunkle LM, Kotloff KL, Gay CL, et al; 2019nCoV-301 Study Group. Efficacy and safety of NVX-CoV2373 in adults in the United States and Mexico. N Engl J Med. 2022;386(6):531–543.

15. Heath PT, Galiza EP, Baxter DN, et al; 2019nCoV-302 Study Group. Safety and efficacy of the NVX-CoV2373 COVID-19 vaccine. N Engl J Med. 2021;385(13):1172–1183.

16. Bennett C, Rivers EJ, Woo W, Bloch M, Cheung K, Griffin P, et al. Immunogenicity and safety of heterologous Omicron BA.1 and bivalent SARS-CoV-2 recombinant spike protein booster vaccines: a phase 3, randomized, clinical trial. J Infect Dis. 2023 Nov 16:jiad508.

17. Koutsakos M, Ellebedy AH. Immunological imprinting: Understanding COVID-19. Immunity. 2023;56(5):909–913. doi: 10.1016/j.immuni.2023.04.012. Epub 2023 Apr 19. PMID: 37105169; PMCID: PMC10113596.

